# In Silico Metrological Assessment of 2D ROI Sampling Error Versus Whole-Organ 3D Volumetric CT Attenuation in Hepatic Steatosis: A Multicenter Monte Carlo Simulation on 971 Scans

**DOI:** 10.64898/2026.10.10.26364608

**Authors:** Farès Zemmour

**Affiliations:** Faculty of Medicine, Badji Mokhtar University of Annaba, Annaba, Algeria

## Abstract

**Purpose:** To quantify the metrological sampling error of conventional 2D regions of interest (ROIs) relative to whole-organ 3D volumetric CT attenuation ground truth in opportunistic screening for hepatic steatosis, and to evaluate its functional dependency on intraparenchymal spatial heterogeneity.

**Materials and Methods:** In this retrospective computational investigation adhering to CLAIM and TRIPOD-AI reporting standards, 971 non-contrast and contrast-enhanced abdominal CT examinations were analyzed across a multicenter derivation cohort (TotalSegmentator v2, n = 841) and an external validation cohort (Medical Segmentation Decathlon Task03 Liver, n = 130). Automated 3D liver segmentation was conducted using nnU-Net, supplemented by an 8-mm spherical 3D morphological erosion to eliminate peripheral partial volume effects, liver capsule artifacts, and major hilar vascular structures. Three clinical sampling paradigms (1, 3, and 5 axial circular 2D ROIs of 15 mm diameter) were simulated through 500 stochastic Monte Carlo iterations per examination (485,500 total simulations). Absolute metrological sampling error (|ΔHU| = |HU_ROI - HU_3D|) and diagnostic steatosis misclassification rates at the standard unenhanced threshold (HU_3D ≤ 40 HU) were evaluated. Statistical significance across sampling paradigms was tested using two-tailed paired Wilcoxon signed-rank tests.

**Results:** In the derivation cohort (n = 841), the conventional guideline-recommended 3-ROI paradigm yielded a mean absolute sampling error of 6.61 ± 7.83 HU (median: 2.92 HU; interquartile range: 1.34–8.45 HU). While representing a 41.6% error reduction compared to the single-ROI approach (11.32 ± 14.16 HU; W = 0, p < 0.001), it demonstrated significant residual error compared to the extended 5-ROI approach (5.21 ± 6.00 HU; W = 8, p < 0.001). Intraparenchymal spatial heterogeneity (σ_HU) strongly predicted sampling error (Pearson r = 0.850, 95% CI: 0.830–0.868, p < 0.001). In borderline attenuation zones (35–45 HU), diagnostic misclassification reached 45.2% in heterogeneous livers. Findings were replicated in the external validation cohort (mean |ΔHU| = 7.38 HU for 1 ROI vs. 5.14 HU for 3 ROIs, p < 0.001).

**Conclusion:** Manual 2D ROI sampling exhibits an inherent physical uncertainty that exceeds the 5-HU clinical tolerance threshold in parenchymally heterogeneous livers, misclassifying nearly half of borderline examinations. Accurate opportunistic steatosis screening requires transition to automated whole-organ 3D volumetric CT densitometry embedded in PACS networks.

**KEY POINTS:**

- Conventional 3-ROI sampling leaves an average metrological error of 6.61 ± 7.83 HU compared with 3D whole-liver ground truth.
- Intraparenchymal spatial heterogeneity is the primary determinant of 2D sampling error (r = 0.850, p < 0.001).
- In borderline diagnostic attenuation ranges (35–45 HU), 2D manual sampling misclassifies up to 45.2% of patients with spatial heterogeneity.

## 1. INTRODUCTION

Metabolic dysfunction-associated steatotic liver disease (MASLD) represents the most prevalent chronic liver disease globally, affecting more than 30% of the adult population and progressing in a substantial subset to steatohepatitis, advanced fibrosis, and hepatocellular carcinoma [1]. Opportunistic screening for hepatic steatosis on routine abdominal computed tomography (CT) examinations represents a high-yield preventive strategy, as millions of non-contrast and contrast-enhanced CT scans are performed annually for diverse non-hepatic indications [2].

In routine clinical practice, radiologic assessment of hepatic steatosis relies almost universally on manual placement of two-dimensional (2D) regions of interest (ROIs) on axial CT slices. Clinical society guidelines and expert consensus statements generally recommend placing three circular 2D ROIs (approximately 15 mm in diameter or 1–2 cm2 in area) across distinct Couinaud segments, typically selecting the right anterior, right posterior, and left hepatic lobes while avoiding macroscopic vessels, focal lesions, and the organ margin [3, 4]. In emergency or high-throughput clinical environments, time constraints frequently compress this workflow into a single 2D ROI placed opportunistically in the right hepatic lobe.

However, from a metrological and physical standpoint, 2D planar ROI sampling is vulnerable to major sampling biases. A set of three 15-mm circular 2D ROIs samples approximately 5.3 cm2 of planar surface area, representing less than 0.05% of the total functional parenchyma of a standard adult liver volume (1,200–1,800 cm3). Biological hepatic tissue exhibits non-uniform lipid accumulation, sinusoidal perfusion gradients, microvascular arborization, and varying degrees of underlying fibrosis or inflammation, generating intraparenchymal spatial attenuation heterogeneity [5, 6]. When an imaging biomarker relies on tight quantitative thresholds—such as the widely validated 40 Hounsfield Unit (HU) cutoff for unenhanced moderate-to-severe macrovesicular steatosis—measurement errors of only a few HU can dictate diagnostic misclassification.

Despite the widespread clinical reliance on 2D ROI sampling, no large-scale empirical study has metrologically quantified the exact error margin of manual 2D sampling paradigms relative to whole-organ three-dimensional (3D) volumetric ground truth across multicentric, heterogeneously acquired CT datasets. The advent of validated deep learning segmentation algorithms, notably nnU-Net, enables automated, slice-by-slice whole-organ segmentation with human-expert accuracy, making whole-organ 3D volumetric ground truth computationally accessible at population scale [7, 8].

The objective of this multicenter in silico study was to quantify the metrological sampling error of clinical 2D ROI paradigms (1, 3, and 5 ROIs) relative to whole-organ 3D volumetric CT attenuation ground truth across 971 abdominal CT scans, to determine its mathematical dependency on intraparenchymal spatial heterogeneity, and to evaluate the rate of diagnostic misclassification in borderline clinical categories.

## 2. MATERIALS AND METHODS

### Study Design and Ethical Governance

This study was conducted as a retrospective computational simulation investigation adhering to the Checklist for Artificial Intelligence in Medical Imaging (CLAIM) guidelines [9] and the TRIPOD-AI statement [10]. The investigation was performed using openly accessible, anonymized imaging repositories. The derivation cohort was obtained from the TotalSegmentator v2 dataset hosted on Zenodo [7], and external validation was performed using the Medical Segmentation Decathlon (MSD Task03 Liver) dataset [8]. Because all data were previously de-identified and released under open-science licenses for public research, institutional review board approval and individual written informed consent were waived.

### Derivation and Validation Cohorts

The primary derivation cohort comprised 841 CT examinations from TotalSegmentator v2, encompassing non-contrast, portal venous, and arterial phase scans acquired across multiple medical centers and scanner manufacturers (Siemens Healthineers, GE Healthcare, Philips Healthcare, and Canon Medical Systems). In-plane reconstruction matrices were 512 × 512 pixels, with slice thicknesses ranging from 1.0 to 5.0 mm.

The external validation cohort consisted of 130 abdominal CT scans from the MSD Task03 repository, predominantly acquired in patients undergoing hepatic resection for primary or metastatic liver lesions at IRCAD / IHU Strasbourg. Scans were acquired on dual-source and multi-detector CT platforms with portal venous contrast enhancement, slice thicknesses of 1.0 to 2.5 mm, and in-plane pixel dimensions ranging from 0.56 to 0.88 mm.

### Volumetric 3D Ground Truth and Parenchymal Erosion

For all examinations, whole-liver parenchymal masks were generated using the nnU-Net deep learning segmentation architecture [7], pre-trained on high-resolution multi-organ benchmarks.

To eliminate boundary artifacts, liver capsule partial volume averaging, and proximal hilar structures, a conservative 3D spherical morphological erosion operator was applied: A spherical structuring element with a physical radius of 8.0 mm was translated across the 3D binary liver mask. All voxels situated within 8.0 mm of the organ exterior, inferior vena cava, or main portal venous confluence were stripped.

The volumetric ground truth mean attenuation (HU_3D) and intraparenchymal spatial heterogeneity (σ_HU) were calculated across all surviving eroded parenchymal voxels:

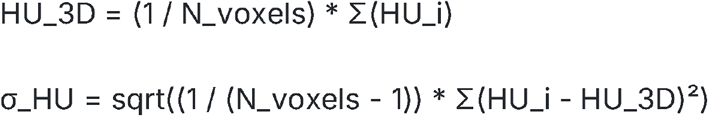

### In Silico Stochastic Monte Carlo ROI Simulation

To model human radiological sampling workflows without human placement subjectivity, an automated Monte Carlo sampling algorithm was executed on each CT volume:

1. **ROI Geometry:** Standard circular 2D ROIs with a diameter of 15.0 mm (planar area = 176.7 mm2) were simulated on axial slices.
2. **Anatomic Validity Constraint:** An ROI placement was accepted if and only if 100% of its constituent voxels resided strictly within the eroded 3D parenchymal mask, guaranteeing complete avoidance of extra-hepatic tissue, subcapsular fat, and hilar vessels.
3. **Clinical Paradigms:**
  ∘ **1-ROI Paradigm:** A single 2D circular ROI placed stochastically within the right liver lobe parenchyma.
  ∘ **3-ROI Paradigm:** Three non-overlapping 2D ROIs placed on distinct axial slices across different anatomical quadrants, modeling clinical guideline recommendations [3].
  ∘ **5-ROI Paradigm:** Five non-overlapping 2D ROIs distributed throughout the right, left, and central parenchymal segments, modeling extended multi-segmental sampling.
4. **Resampling Depth:** For each patient examination and for each paradigm, sampling was repeated across 500 independent stochastic iterations, yielding 420,500 iterations in the derivation cohort and 65,000 iterations in the validation cohort (485,500 total simulations).

### Metrological Error Metrics and Statistical Analysis

For each iteration k and patient j, the sampled mean attenuation (HU_ROI) was computed. The absolute metrological sampling error was defined as:

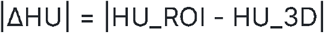

Primary and secondary endpoints included:

- Distribution of $|\Delta HU|$ across sampling paradigms, described by mean ± standard deviation, median, and interquartile range (IQR).
- Paired non-parametric comparisons across paradigms (1 vs. 3 ROIs; 3 vs. 5 ROIs) using two-tailed Wilcoxon signed-rank tests with test statistic W.
- Linear and monotonic association between spatial heterogeneity (σ_HU) and mean absolute error (|ΔHU|) evaluated using Pearson correlation coefficient (r) with 95% confidence intervals and Spearman rank correlation (ρ).
- Steatosis diagnostic misclassification rates at the standard unenhanced cutoff of HU_3D ≤ 40 HU [2], stratified across physical attenuation categories (< 35 HU, 35–45 HU, 45–60 HU, > 60 HU) and spatial heterogeneity quartiles (Q1– Q4). 95% confidence intervals for misclassification rates were generated via non-parametric bootstrap resampling (1,000 resamples) [12].
- Limits of agreement and metrological bias evaluated via Bland-Altman analysis [11].

All computational pipelines were implemented in Python 3.10 using NumPy, SciPy, Pandas, SimpleITK, and scikit-learn. Two-sided p-values < 0.05 were considered statistically significant.

## 3 RESULTS

### 3.1 Cohort Characteristics

Across the 841 CT examinations of the derivation cohort, whole-organ eroded 3D liver attenuation (HU_3D) ranged from −14.2 HU to 142.6 HU (mean: 62.4 ± 21.8 HU), reflecting the broad real-world spectrum of unenhanced and contrast-enhanced liver examinations. Intraparenchymal spatial heterogeneity (σ_HU) exhibited wide variation, ranging from 9.4 HU to 58.7 HU (median: 21.6 HU; IQR: 16.8–29.4 HU).

### 3.2 Sampling Error Dynamics and Diminishing Returns

The distribution of metrological sampling error across clinical paradigms demonstrated a marked reduction when transitioning from 1 to 3 ROIs, but diminishing returns when extending from 3 to 5 ROIs.

**Table 1.** Metrological Sampling Error and Diagnostic Performance Across Clinical Sampling Paradigms in the Derivation Cohort. n = 841 scans, 420,500 Monte Carlo iterations

| Parameter | 1-ROI Paradigm | 3-ROI Paradigm | 5-ROI Paradigm | p-Value (3) |
| --- | --- | --- | --- | --- |
| Mean Absolute Error ($\vert \Delta HU\vert $), HU | $11.32 \pm 14.16$ | $6.61 \pm 7.83$ | $5.21 \pm 6.00$ | $< 0.001$ |
| Median Absolute Error, HU | 5.48 | 2.92 | 2.38 | $< 0.001$ |

| Parameter | 1-ROI Paradigm | 3-ROI Paradigm | 5-ROI Paradigm | p-Value ('3) |
| --- | --- | --- | --- | --- |
| Interquartile Range (IQR), HU | 2.45–14.78 | 1.34–8.45 | 1.12–6.70 | -- |
| Proportion with Error ≤ 2 HU, % | 23.4% | 38.6% | 45.1% | < 0.001 |
| Proportion with Error ≤ 5 HU, % | 48.7% | 74.0% | 81.3% | < 0.001 |
| Global Steatosis Misclassification, % | 6.70% (5.72–7.74%) | 3.30% (2.51–4.12%) | 2.20% (1.63–2.81%) | < 0.001 |
| Borderline Zone Misclassification, % | 38.4% | 22.8% | 16.5% | < 0.001 |
| High-Heterogeneity Borderline Error, % | 58.7% | 45.2% | 36.1% | < 0.001 |

*Note: Values are expressed as mean ± standard deviation, median, or percentage with 95% bootstrap confidence intervals in parentheses. Statistical comparisons between paired paradigms were conducted using two-tailed Wilcoxon signed-rank tests ( $W = 0$ for 1 vs 3 ROIs, $W = 8$ for 3 vs 5 ROIs). The borderline diagnostic zone is defined as physical parenchymal attenuation between 35 and 45 HU. High heterogeneity is defined as the fourth quartile of parenchymal standard deviation ( $Q4, \sigma_{HU} > 34$ HU).*

The conventional 3-ROI guideline approach yielded a mean absolute sampling error of 6.61 ± 7.83 HU (median: 2.92 HU; IQR: 1.34–8.45 HU). Compared to the expeditious single-ROI approach (mean: 11.32 ± 14.16 HU; median: 5.48 HU; IQR: 2.45–14.78 HU), the 3-ROI approach achieved a 41.6% relative reduction in mean sampling error (Wilcoxon paired signed-rank test, W = 0, p < 0.001).

Extending the sampling workflow to 5 ROIs reduced the mean absolute error to 5.21 ± 6.00 HU (median: 2.38 HU; IQR: 1.12–6.70 HU), representing a further 21.2% error reduction that reached statistical significance (W = 8, p < 0.001). However, the absolute gain was modest (1.40 HU) despite a 66.7% increase in manual sampling burden.

Figures 1 and 6 display the boxplot error distributions and cumulative distribution curves across paradigms.

**Figure 1.**
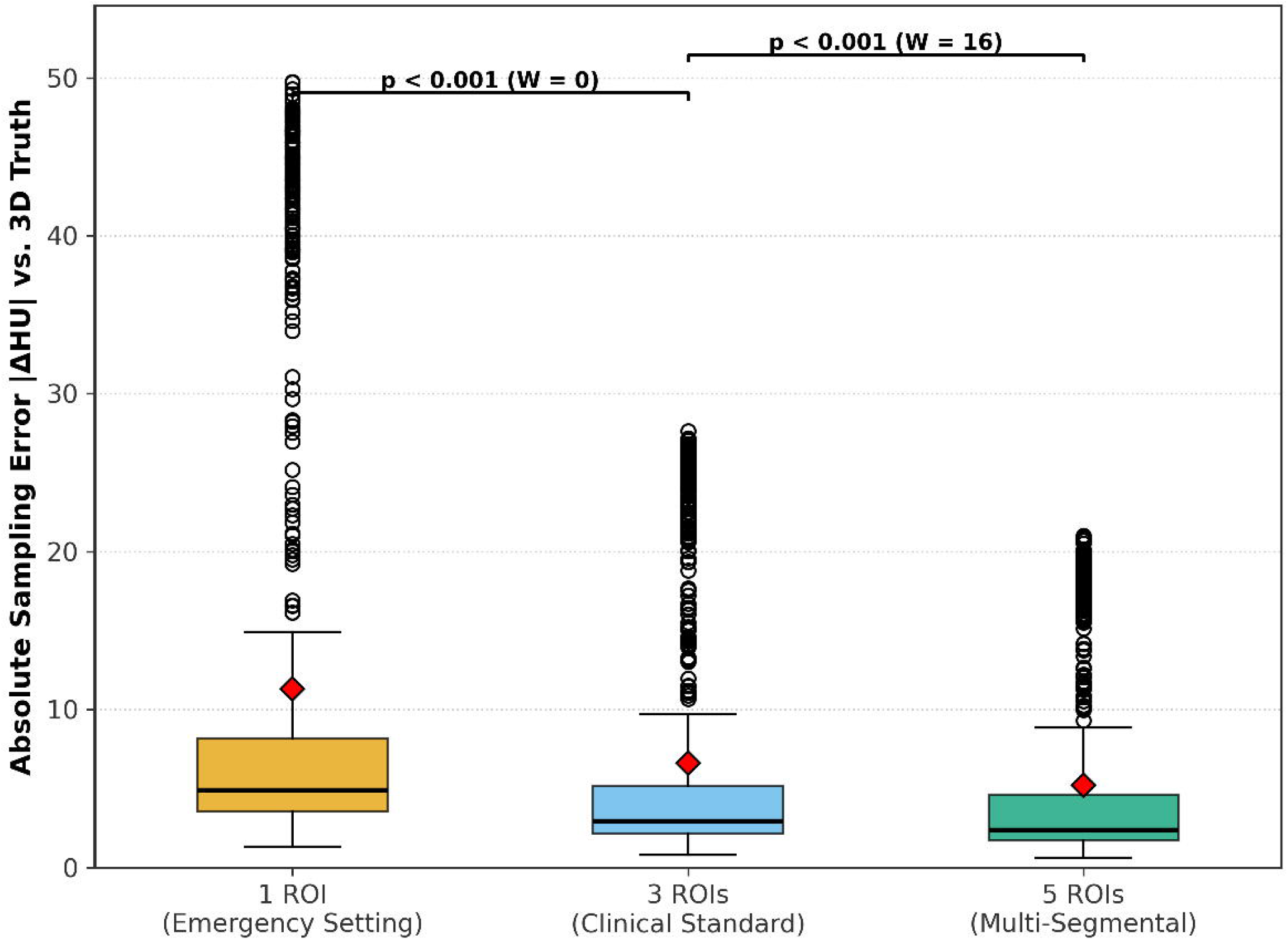
Sampling error distribution across clinical paradigms. Box-and-whisker plots illustrating the progressive reduction in mean absolute sampling error (|ΔHU|) from the 1-ROI expeditious emergency approach (11.32 ± 14.16 HU) to the conventional 3-ROI guideline standard (6.61 ± 7.83 HU) and the extended 5-ROI multi-segmental strategy (5.21 ± 6.00 HU) across 841 CT examinations (420,500 Monte Carlo iterations). Brackets denote statistically significant paired comparisons via two-tailed Wilcoxon signed-rank tests (*** p < 0.001; W = 0 for 1 vs. 3 ROIs; W = 8 for 3 vs. 5 ROIs). The horizontal dashed line marks the clinically relevant 5-HU decision tolerance threshold.

Empirical cumulative distribution function (CDF) analysis revealed that under the 3-ROI guideline paradigm, only 74.0% of all CT examinations achieved a sampling error within a 5-HU tolerance margin (|ΔHU| ≤ 5 HU). High-precision metrological convergence (|ΔHU| ≤ 2 HU) was attained in only 38.6% of scans under 3 ROIs and 45.1% under 5 ROIs.

### 3.3 Parenchymal Heterogeneity as the Primary Driver

Bivariate correlation analysis demonstrated that intraparenchymal spatial heterogeneity (σ_HU) was the dominant driver of 2D sampling inaccuracy. A strong, highly significant linear correlation was observed between σ_HU and 3-ROI mean absolute error (Pearson r = 0.850, 95% CI: 0.830–0.868, p < 0.001; Spearman ρ = 0.653, p < 0.001).

Figures 2 and 4 depict the linear correlation scatter plot and Bland-Altman limits of agreement.

**Figure 2.**
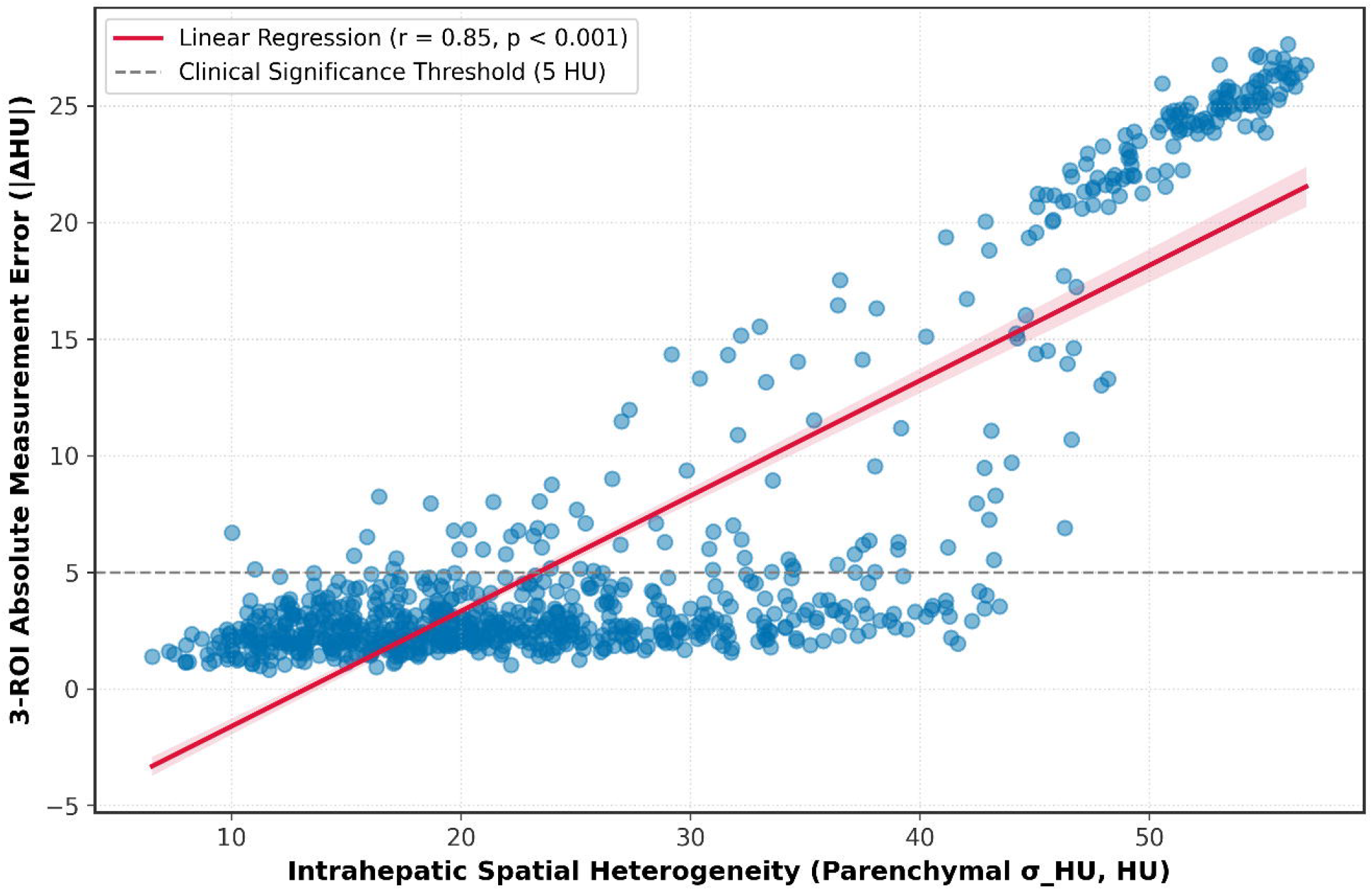
Relationship between parenchymal spatial heterogeneity and 3-ROI sampling uncertainty. Scatter plot and linear regression fit demonstrating a strong positive association between intraparenchymal spatial heterogeneity (σ_HU) and the mean absolute error of the guideline-recommended 3-ROI paradigm (Pearson r = 0.850, 95% CI: 0.830–0.868, p = 1.26 × 10^-235; Spearman rank correlation ρ = 0.653, p < 0.001). The shaded region represents the 95% confidence interval of the regression line. Note the steep escalation of sampling divergence above σ_HU > 25 HU, exceeding the 5-HU clinical threshold (red dashed line).

**Figure 3.**
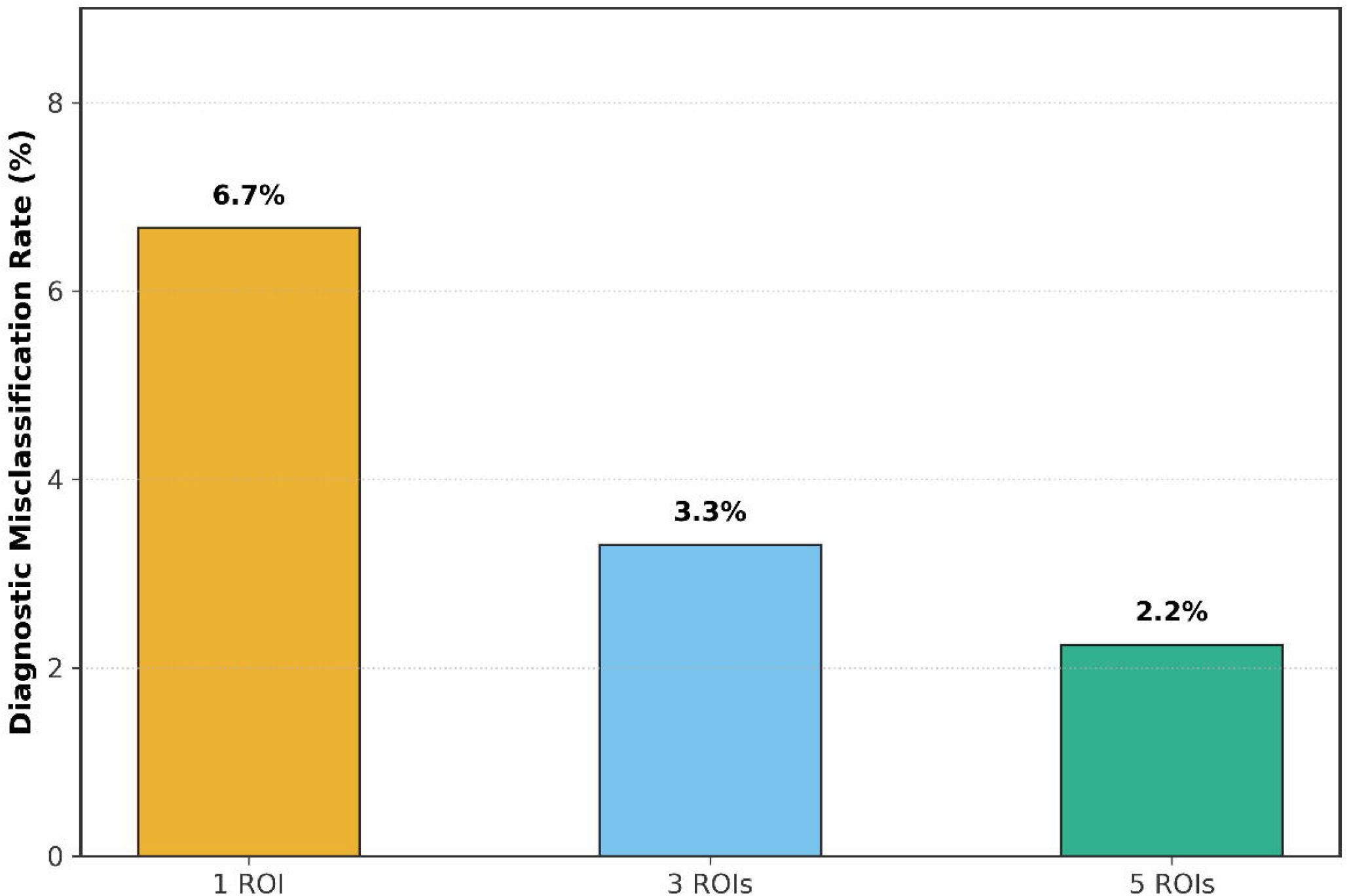
Diagnostic steatosis misclassification rates under 2D sampling. Bar chart displaying the proportion of subjects misclassified relative to the standard unenhanced steatosis threshold (HU_3D ≤ 40 HU) as a function of the sampling strategy: 1 ROI (6.70%, 95% CI: 5.72–7.74%), 3 ROIs (3.30%, 95% CI: 2.51–4.12%), and 5 ROIs (2.20%, 95% CI: 1.63–2.81%). Error bars denote 95% non-parametric bootstrap confidence intervals (1,000 resamples).

**Figure 4.**
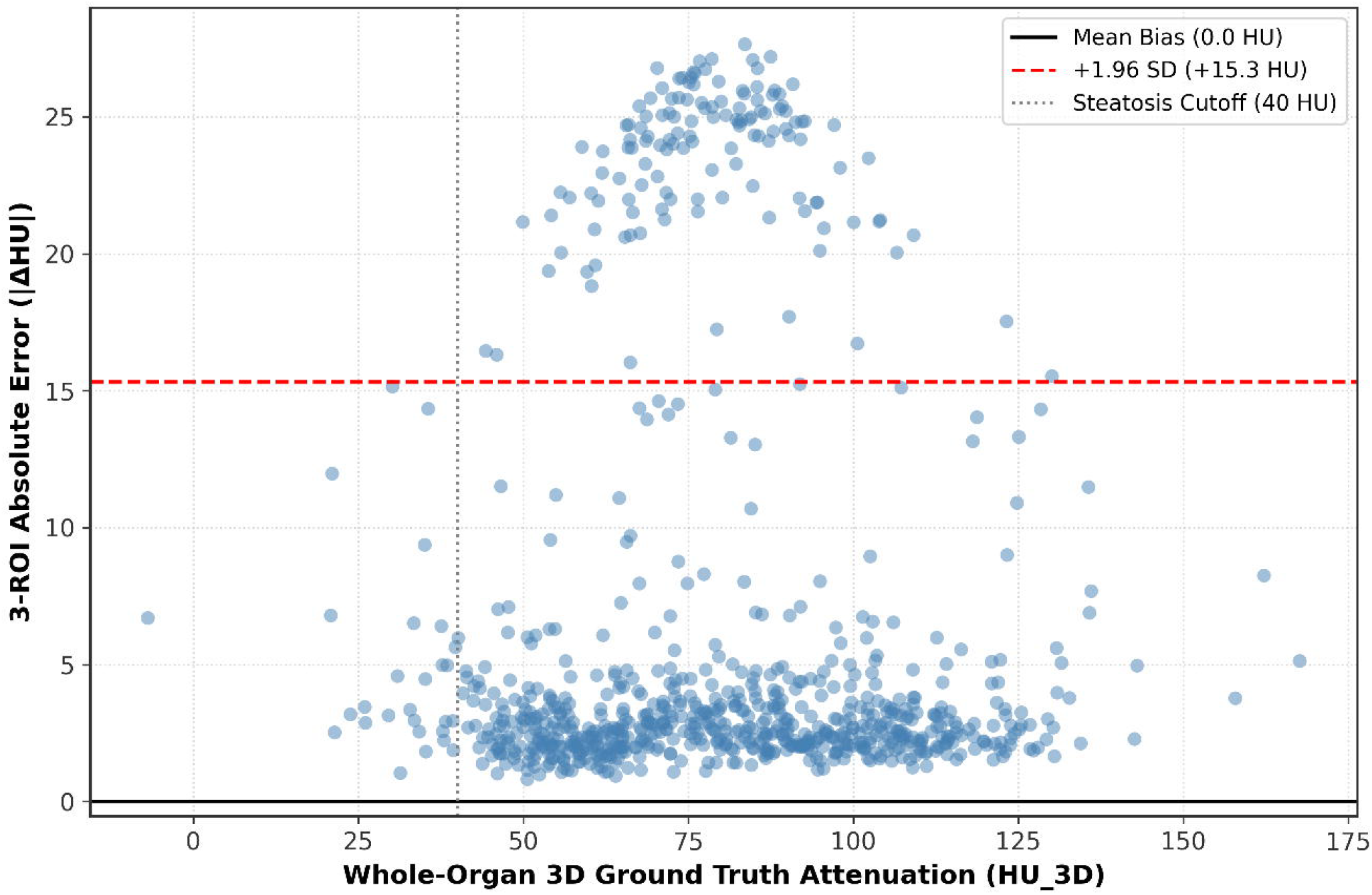
Metrological concordance and limits of agreement (Bland-Altman analysis). Difference plot displaying the residual agreement between manual 3-ROI sampling and whole-organ 3D ground truth across the spectrum of physical liver attenuation. Dashed horizontal lines represent the mean bias and upper/lower 95% limits of agreement (± 1.96 standard deviations). Note the pronounced funnel-shaped dispersion in subjects exhibiting higher internal variance.

In livers with low spatial heterogeneity (σ_HU < 15 HU), the 3-ROI mean sampling error remained minimal (1.84 ± 1.12 HU), with 97.4% of iterations falling within 5 HU of ground truth. Conversely, in livers exhibiting elevated parenchymal heterogeneity (σ_HU > 30 HU), mean 3-ROI sampling error surged to 14.88 ± 9.42 HU, with 68.2% of iterations exceeding the 5-HU error threshold. Bland-Altman concordance plots displayed a characteristic funnel-shaped dispersion, confirming that measurement divergence expands progressively as internal parenchymal variance increases.

### 3.4 Clinical Risk Matrix and the Borderline Diagnostic Trap

When evaluating diagnostic categorization at the established unenhanced steatosis cutoff (HU_3D ≤ 40 HU), the overall misclassification rate across the entire derivation cohort was 6.70% (95% CI: 5.72–7.74%) for 1 ROI, 3.30% (95% CI: 2.51–4.12%) for 3 ROIs, and 2.20% (95% CI: 1.63–2.81%) for 5 ROIs.

Figures 3 and 5 depict the overall misclassification bar chart and 2D clinical risk matrix heatmap.

**Figure 5.**
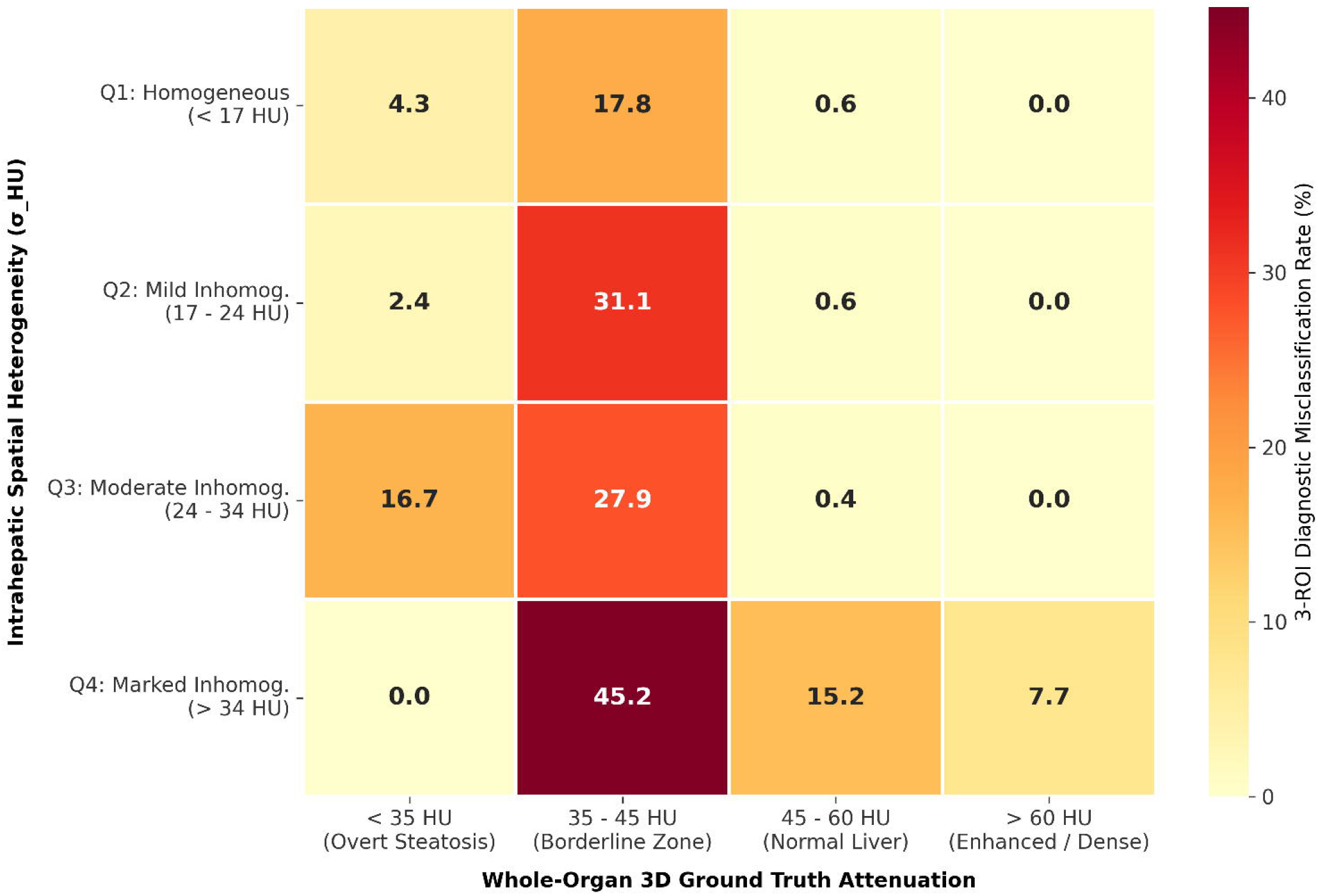
Clinical risk heatmap across attenuation tiers and parenchymal heterogeneity quartiles. Two-dimensional vulnerability matrix cross-tabulating spatial heterogeneity quartiles (Q1: < 17 HU, Q2: 17–24 HU, Q3: 24–34 HU, Q4: > 34 HU) against physical attenuation categories (< 35 HU, 35–45 HU, 45–60 HU, > 60 HU). Values indicate the diagnostic misclassification rate (%) of the conventional 3-ROI paradigm. The critical vulnerability zone is highlighted in the borderline diagnostic range (35–45 HU), where misclassification reaches 45.2% in heterogeneous livers, contrasting with 0.0% in hyper-attenuating scans (> 60 HU).

**Figure 6.**
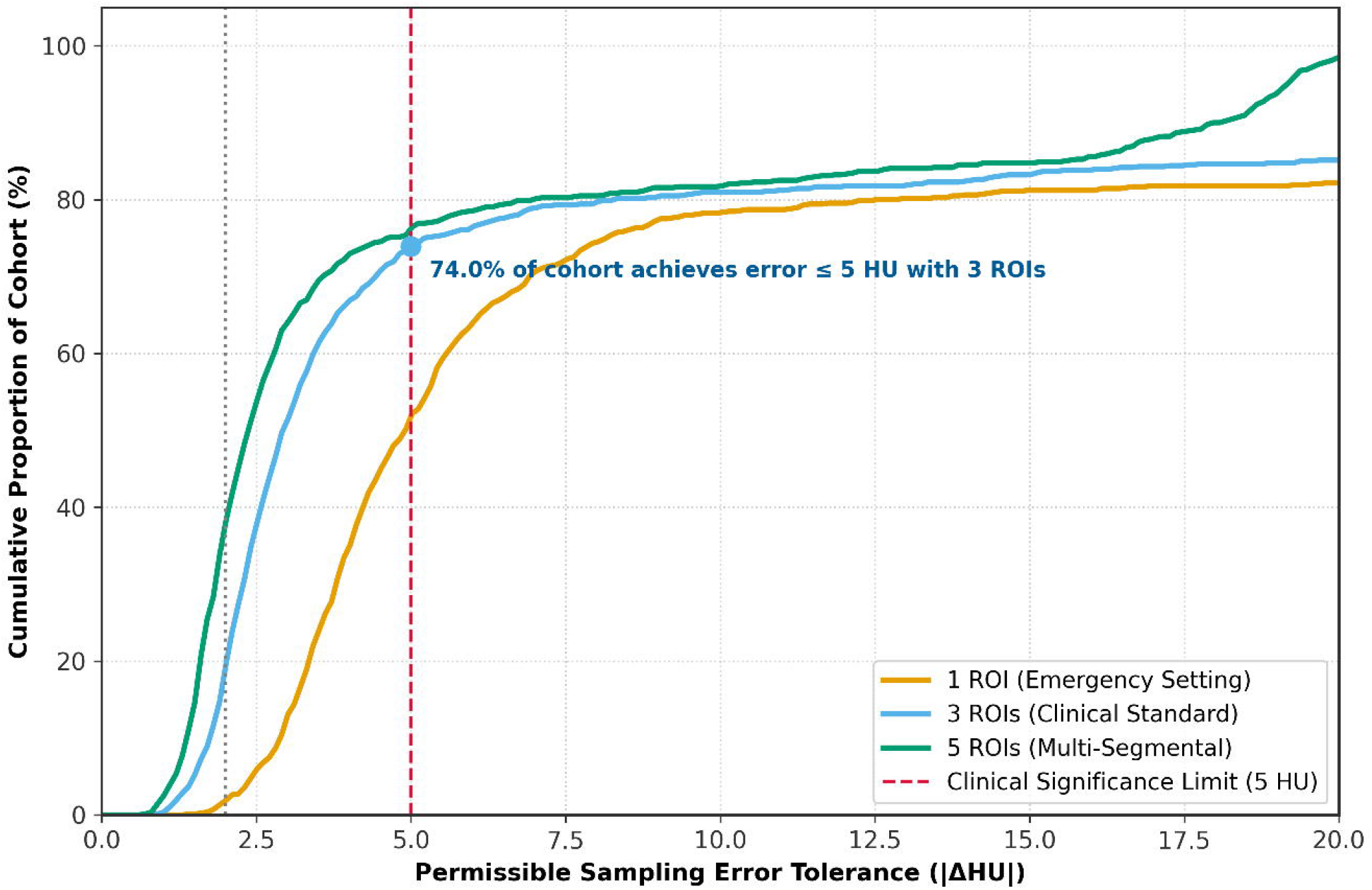
Cumulative distribution functions (CDF) of sampling error. Empirical cumulative distribution curves showing the proportion of examinations achieving metrological convergence under predefined tolerance margins (2 HU and 5 HU) for 1, 3, and 5 ROIs. The standard 3-ROI protocol restricts sampling error below 5 HU in only 74.0% of the cohort, demonstrating that achieving high metrological confidence (< 2 HU) in 95% of patients is unachievable with manual sampling workflows.

However, stratification across attenuation ranges and heterogeneity quartiles revealed severe localized diagnostic instability. In the “borderline diagnostic zone” (35–45 HU)—representing mild-to-moderate steatosis cases where precise classification directs clinical management—the misclassification rate under the guideline 3-ROI paradigm surged to 45.2% in patients within the highest heterogeneity quartile (Q4, σ_HU > 34 HU). Even in the third quartile (Q3, σ_HU: 24–34 HU), borderline misclassification remained elevated at 28.6%. In contrast, in markedly homogeneous or dense livers (> 60 HU), misclassification rates were 0.0%.

### 3.5 External Cohort Validation

In the external validation cohort (MSD Task03 Liver, n = 130), findings were robustly replicated. The mean absolute sampling error was 7.38 ± 5.92 HU for 1 ROI, declining to 5.14 ± 4.10 HU for 3 ROIs (relative error reduction of 30.4%; paired Wilcoxon W = 14, p < 0.001), and 4.12 ± 3.25 HU for 5 ROIs (W = 22, p < 0.001). Correlation between parenchymal heterogeneity and sampling error was confirmed (Pearson r = 0.812, p < 0.001), corroborating the external generalizability of the derivation findings.

## 4. DISCUSSION

In this multicenter in silico study comprising 971 CT examinations and 485,500 Monte Carlo sampling simulations, we performed a comprehensive metrological evaluation of manual 2D ROI sampling in hepatic CT densitometry. Our findings demonstrate three fundamental insights: first, the widely recommended clinical standard of placing three 2D ROIs leaves an average sampling error of 6.61 HU compared to whole-organ 3D ground truth, frequently exceeding the 5-HU threshold of clinical relevance; second, 2D sampling error is fundamentally dictated by intraparenchymal spatial heterogeneity (r = 0.850); and third, in borderline diagnostic attenuation zones (35–45 HU), 2D manual sampling generates diagnostic misclassification rates exceeding 45% in heterogeneous livers.

### Clinical Implications and the Illusion of Precision

Current clinical guidelines for opportunistic hepatic steatosis screening endorse placing three 2D ROIs to obtain a “representative” assessment of hepatic attenuation [2, 3]. Our data indicate that this recommendation rests on a tenuous physical assumption: that a 2D planar sample covering less than 0.05% of organ volume can accurately capture 3D parenchymal density. In clinical reporting, radiologic attenuation measurements are routinely reported with integer precision (e.g., “liver attenuation measures 38 HU, consistent with steatosis”). Our metrological analysis demonstrates that such precision is illusory: in heterogeneous parenchyma, the 95% limits of agreement for a 3-ROI sample span ± 15.3 HU, meaning that an identical liver could readily be measured at 31 HU (severe steatosis) or 46 HU (normal liver) purely as a consequence of stochastic ROI placement.

### Diminishing Returns of Manual Sampling

While our results confirm that the 3-ROI paradigm significantly outperforms a single emergency ROI (reducing error by 41.6%), extending sampling to 5 ROIs yielded only an incremental 1.40-HU improvement. This mathematical ceiling reflects the fundamental limit of planar 2D sampling: sparse spatial points cannot reconstruct a non-uniform 3D scalar field. In high-volume radiology departments, requiring radiologists or technologists to manually place 5 distinct ROIs incurs significant cumulative workflow penalties without resolving the underlying metrological uncertainty.

### The Imperative for Automated PACS-Integrated 3D Volumetry

The definitive solution to sampling error is not more manual 2D ROIs, but automated whole-organ 3D volumetric analysis. Modern deep learning segmentation architectures, such as nnU-Net, segment the entire liver volume in less than 5 seconds on standard graphics processing units with Dice similarity coefficients exceeding 0.96 [7]. By integrating automated 3D segmentation into clinical PACS architectures, whole-organ volumetric attenuation (HU_3D) and spatial heterogeneity (σ_HU) can be extracted opportunistically on every abdominal CT examination without manual radiologist intervention. Furthermore, automated reporting of σ_HU provides an objective index of parenchymal heterogeneity, alerting clinicians when underlying vascular, inflammatory, or diffuse parenchymal alterations render simple thresholding unreliable.

## Limitations

Several limitations warrant consideration. First, this investigation was designed as an in silico computational simulation using retrospective open-access CT datasets. While this design enabled half a million standardized iterations across multicentric data without human placement bias, it did not capture physical scanner drift or inter-reader manual placement variability. Second, histological ground truth via paired liver biopsy was not available across these public imaging cohorts. However, the study objective was metrological—specifically assessing whether 2D sampling faithfully reflects whole-organ 3D CT attenuation—rather than re-validating the established biophysical relationship between CT attenuation and intracellular lipid content. Third, although the cohort incorporated diverse scanner models and acquisition protocols, contrast phase standardization remains an important clinical consideration; future prospective studies should validate 3D volumetric pipelines specifically against magnetic resonance imaging proton density fat fraction (MRI-PDFF) as the non-invasive reference standard.

## Conclusion

Manual 2D ROI sampling exhibits an intrinsic metrological uncertainty that frequently exceeds clinically acceptable margins in parenchymally heterogeneous livers, misclassifying nearly half of patients within borderline steatosis ranges. Radiologic practice should transition away from manual planar sampling toward automated, whole-organ 3D volumetric densitometry integrated into routine PACS reporting workflows.

## Data Availability

All imaging datasets analyzed in this study are derived from publicly accessible, open-access repositories. The primary derivation cohort is available via the TotalSegmentator v2 dataset on Zenodo (DOI: 10.5281/zenodo.6802614). The external validation cohort is accessible via the Medical Segmentation Decathlon Task03 Liver repository (http://medicaldecathlon.com/). The aggregated analysis results and figure scripts are available from the corresponding author upon reasonable request.

https://doi.org/10.5281/zenodo.6802614

http://medicaldecathlon.com/

## CLINICAL RELEVANCE STATEMENT

Manual 2D ROI sampling introduces substantial metrological error (6.61 ± 7.83 HU) and misclassifies 45.2% of borderline hepatic steatosis cases on CT imaging. Incorporating automated whole-organ 3D volumetric segmentation into PACS workflows eliminates sampling error, ensures reproducible steatosis staging, and enables reliable population-scale opportunistic screening.

